# Leadership, Informatics Expertise, and Resources: Determinants of Institutional Data Sharing in the National Clinical Cohort Collaborative (N3C)

**DOI:** 10.1101/2025.05.01.25326202

**Authors:** Carly M. Rose, William S. Bush, Mark F. Beno, Scott M. Williams, Jonathan L. Haines, Dana C. Crawford

**Affiliations:** Program in Biomedical & Health Informatics; Cleveland Institute for Computational Biology; Department of Population and Quantitative Health Sciences; Department of Genetics and Genome Sciences Case Western Reserve University, Cleveland, OH

## Abstract

The National Clinical Cohort Collaborative (N3C), initially developed in response to the SARS-CoV-2/COVID-19 pandemic, is one of the largest centralized, individual-level clinical data repositories available for research in the US. Some institutions do not share data with N3C. We accessed public data to derive institutional variables representative of leadership, informatics expertise, and resources, potentially addressable factors we hypothesized were associated with N3C clinical data sharing. N3C data sharing institutions were more likely to have active NCATS funding, have affiliated departments, centers, or institutes of biomedical informatics, and active Fellows of the American College of Medical Informatics faculty compared with institutions that did not share data with N3C. While we identified institutional characteristics associated with N3C data sharing, there are likely other characteristics associated with institutional data sharing decisions. Understanding factors that motivate or deter institutional data sharing for research is key to addressing barriers and making more data open.

## Introduction

Electronic health records (EHRs) hold enormous research potential; however, US EHRs are siloed by institution and not easily accessible to a broad range of researchers, limiting timely, population-scale analyses. In response to the COVID-19 pandemic, the National Clinical Cohort Collaborative (N3C), a National Center for Advancing Translational Sciences (NCATS) initiative, addressed this compartmentalization to promote rapid clinical data sharing by collecting, harmonizing, and centralizing EHR data across clinical institutions to further COVID-19 research in the United States^1^. The N3C offers a unique opportunity to access national, de-identified, individual-level EHR data from nearly 100 institutions across the United States for rapid and meaningful research on COVID-19.

As of June 20, 2024, the N3C data enclave housed 33.2 billion rows of clinical data representing 1.8 billion clinical visits for 22.7 million patients, of whom almost 8.9 million were COVID-19 positive cases^2^. N3C offers three levels of data for investigators at institutions with an active data use agreement: 1) limited dataset (requires local Human Research Protection Program (HRPP) IRB determination), 2) de-identified dataset (truncated zip codes; shifted dates of service), and 3) a synthetic dataset. In comparison, the All of Us Research Program, a National Institutes of Health cohort that began ascertainment in 2019^3^, included just over 466,000 participants with EHR data as of April 2, 2025^4^. All of Us also offers three tiers of data access: 1) public tier (aggregated data only), registered tier (individual-level de-identified data), and 3) restricted tier (individual-level de-identified data linked to genomics; previously suppressed demographic data; unshifted dates and events), none of which require local IRB determination. Like traditional cohorts, All of Us approaches individual participants for consent to access their EHRs whereas N3C invites institutions to enter into data use agreements to share their patient EHRs.

While N3C is now one of the largest repositories of centralized, individual-level EHR data available for COVID-19 research, not all US institutions with EHRs contribute data. We hypothesize here that institutional characteristics such as leadership, informatics expertise, and resources influence an institution’s ability or willingness to share EHR data with N3C (Figure 1). Understanding the reasons for not sharing data should enable the development of systems to overcome data silos, thereby promoting better and more generalizable research.

**Figure 1:**
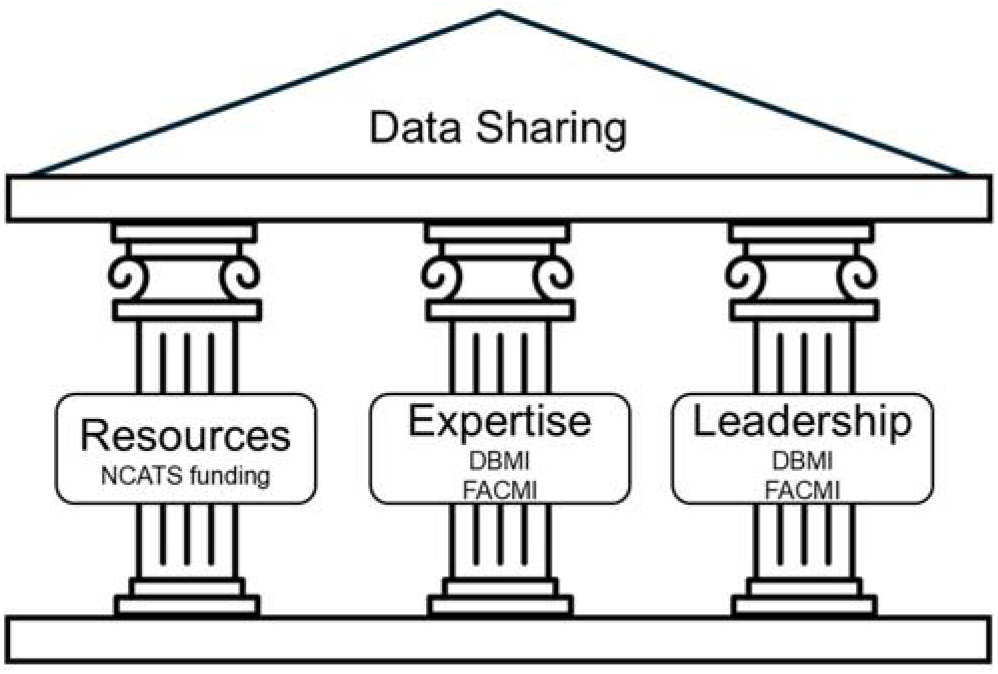
Hypothesized pillars of data sharing. We expect that institutions sharing electronic health record data with the National Clinical Cohort Collaborative (N3C) are strong in informatics resources, expertise, and leadership—three non-mutually exclusive pillars of data sharing examined here. Within each pillar, we mapped the derived variables tested for association with N3C data sharing. Abbreviations: Department of Biomedical Health Informatics (DBMI), Fellow of the American College of Medical Informatics (FACMI), National Center for Advancing Translation Sciences (NCATS).

## Methods

As of July 2022, the N3C had 281 participating institutions: 95 had active Data Transfer Agreements (DTA) and 265 had active Data Use Agreements (DUA). For each participating institution, we accessed publicly available information to define geographic region of the institution (from institutional websites), NCATS funding status (from NIH Reporter^5^), hospital affiliations (from institutional websites), associations with departments, centers, and institutes of biomedical informatics (from institutional websites), and associations with Fellows of the American College of Medical Informatics (FACMI from the American Medical Informatics Association or AMIA website^6^). Geographic region was based on the address of each institution’s main headquarters or campus. Institutions were classified into the US Census Bureau regions of the West, Midwest, South, and Northeast based on zip codes.

Hospital associations were manually examined to determine whether each institution with a DUA but no DTA had clinical data to share with N3C. Institutions included both standalone hospitals and universities with associated hospitals. University health centers or loosely affiliated healthcare providers were not included. For example, the University of Utah has clinical data relevant to N3C (DTA possible) as it is associated with the University of Utah hospital as part of the University of Utah Health healthcare system. In contrast, while Case Western Reserve University is academically affiliated with several Cleveland area hospital systems, they (Cleveland Clinic, University Hospitals, MetroHealth System, and Louis Stokes Cleveland Department of Veterans Affairs Medical Center as part of a collaborative), all are separate institutions requiring separate DUAs. Of these northeast Ohio institutions, Case Western Reserve University does not have its own clinical data to share (DTA not possible) whereas the others do (DTA possible). Institutions with an N3C DTA were classified as an institution with clinical data to share. Becker’s Hospital Review “100 of the largest hospitals and health systems in America 2023” was accessed to identify large hospitals with EHRs not participating in N3C.

To assess institutional capacity and interest in sharing data, we recorded whether individual institutions considered here had a department of biomedical informatics (DBMI) or faculty designated as FACMI. We hypothesized that institutions with DBMIs or FACMI faculty would be more likely and willing to contribute data to large databases like N3C compared with institutions without DBMIs or FACMI faculty. DBMIs status required a department but allowed for slight variations in name, including department of biomedical and health informatics, department of biohealth informatics, and department of biomedical informatics and medical education. One institution had a school of biomedical informatics and was included in the DBMI category given that having a school dedicated to biomedical informatics is evidence of substantial institutional resources dedicated to that field.

Centers and institutes for biomedical informatics (variants of DBMI) were also captured for a sensitivity analysis, but biomedical informatics and related graduate programs administered by non-DBMIs were not. AMIA’s FACMI directory^6^ was accessed to map individual FACMI to the N3C participating institutions. Current FACMI through the class of 2022 were included, excluding retired or deceased FACMI.

Data were input into the REDCap platform, and tests of association were performed using contingency tables and logistic regression using R version 4.2.2. Basic comparisons by DUA/DTA status were made using Fisher’s exact tests of association for categorical variables, and Kruskal-Wallis tests of association for continuous variables, which focused on having a DTA as the outcome of interest. Correlations between independent variables showed no evidence of multicollinearity. The highest variance inflation factor (VIF), calculated using the R *car* package, was 1.10. Since a VIF of 1 represents no correlation and values below 5 are generally considered acceptable, this indicates no problematic multicollinearity. Tests of association were limited to N3C participating institutions with hospital associations (n=126) and 11 large hospitals from the Becker’s Hospital Review 2023 not participating in N3C. The 137 institutions with potential to share clinical data with N3C included 77 with DTA & DUA, 16 with DTA only, 33 with DUA only, and 11 with neither (Table 1; “A Hospital Affiliation” row). We performed univariate analyses with data sharing or DTA (yes/no) as the dichotomous outcome and tested its association with each independent variable (current NCATS funding, affiliated DBMI, variants of DBMI, at least one active FACMI, and number of FACMI per institution). The fully adjusted models included all three independent variables with DTA status as the outcome: current NCATS funding, affiliated DBMI, and at least one active FACMI (Model 1), current NCATS funding, variants of DBMI, and at least one active FACMI (Model 2), current NCATS funding, variants of DBMI, and number of FACMI per institution (Model 3), and current NCATS funding, affiliated DBMI, and number of FACMI per institution (Model 4).

**Table 1.** Institutional Characteristics, by N3C data use (DUA) and data transfer (DTA) agreement status.

|  | DTA & DUA<br>n = 79 | DTA only<br>n = 16 | DUA only<br>n = 186 | Neither DTA<br>nor DUA<br>n = 11 | P-value |
| --- | --- | --- | --- | --- | --- |
| **West** | 15 (19.0%) | 2 (12.5%) | 34 (18.5%) | - | 0.227 |
| **Midwest** | 21 (26.6%) | 3 (18.8%) | 28 (15.2%) | 4 (36.4%) |  |
| **South** | 27 (34.2%) | 7 (43.8%) | 82 (44.6%) | 7 (63.6%) |  |
| **Northeast** | 16 (20.3%) | 4 (25.0%) | 40 (21.7%) | - |  |
| **US Territory** | - | - | 2 (1.1%) | - |  |
| **Current NCATS Funding,**<br>**n (%)** | 31 (39.2%) | 1 (6.3%) | 22 (9.7%) | 1 (9.09%) | <0.001 |
| **A Hospital Affiliation, n**<br>**(%)** | 77 (97.5%) | 16 (100%) | 33 (17.7%) | 11 (100%) | <0.001 |
| **An Affiliated DBMI, n**<br>**(%)** | 12 (15.2%) | 2 (13.3%) | 5 (4.1%) | - | 0.001 |
| **An Affiliated DBMI or**<br>**Centers and Institutes of**<br>**BMI, n (%)** | 41 (51.9%) | 4 (25.0%) | 18 (9.7%) | - | <0.001 |
| **At Least One FACMI, n**<br>**(%)** | 25 (31.65%) | 1 (6.25%) | 18 (9.68%) | - | <0.001 |
| **FACMI per Institution,**<br>**mean (range)** | 1.39 [0-19] | 0.13 [0-2] | 0.27 [0-10] | - | <0.001 |
| *Contingency tables were used to calculate Fisher’s exact p-values for US regions (4x5), Current NCATS Funding (4x2), Hospital Affiliation (4x2), Affiliated DBMI (4x2), and At Least One FACMI (4x2). Kruskal test was used to calculate the p-value for FACMI per institution. US territory institutions are in Puerto Rico.* |  |  |  |  |  |

## Results

Institutions participating in N3C as of mid-2022 spanned all four US geographic regions and Puerto Rico (Table 1 and Figure 2). A plurality (41%) of participating institutions were in the South (based on US Census Bureau definitions), which includes Washington, DC. Among the US institutions participating in N3C (n=281), there were differences by proportion of those: with current NCATS funding (Fisher’s p<0.001); associated with a hospital (Fisher’s p<0.001); associated with a DBMI (Fisher’s p=0.001); associated with variants of a DBMI (Fisher’s p<0.001); associated with at least one active FACMI (Fisher’s p<0.001); and associated with the number of FACMI per institution (Kruskal-Wallis’ p<0.001) by the types of N3C data agreement categories (Table 1). Geographic region did not associate with DUA/DTA status (Fisher’s p>0.05; Table 1).

**Figure 2:**
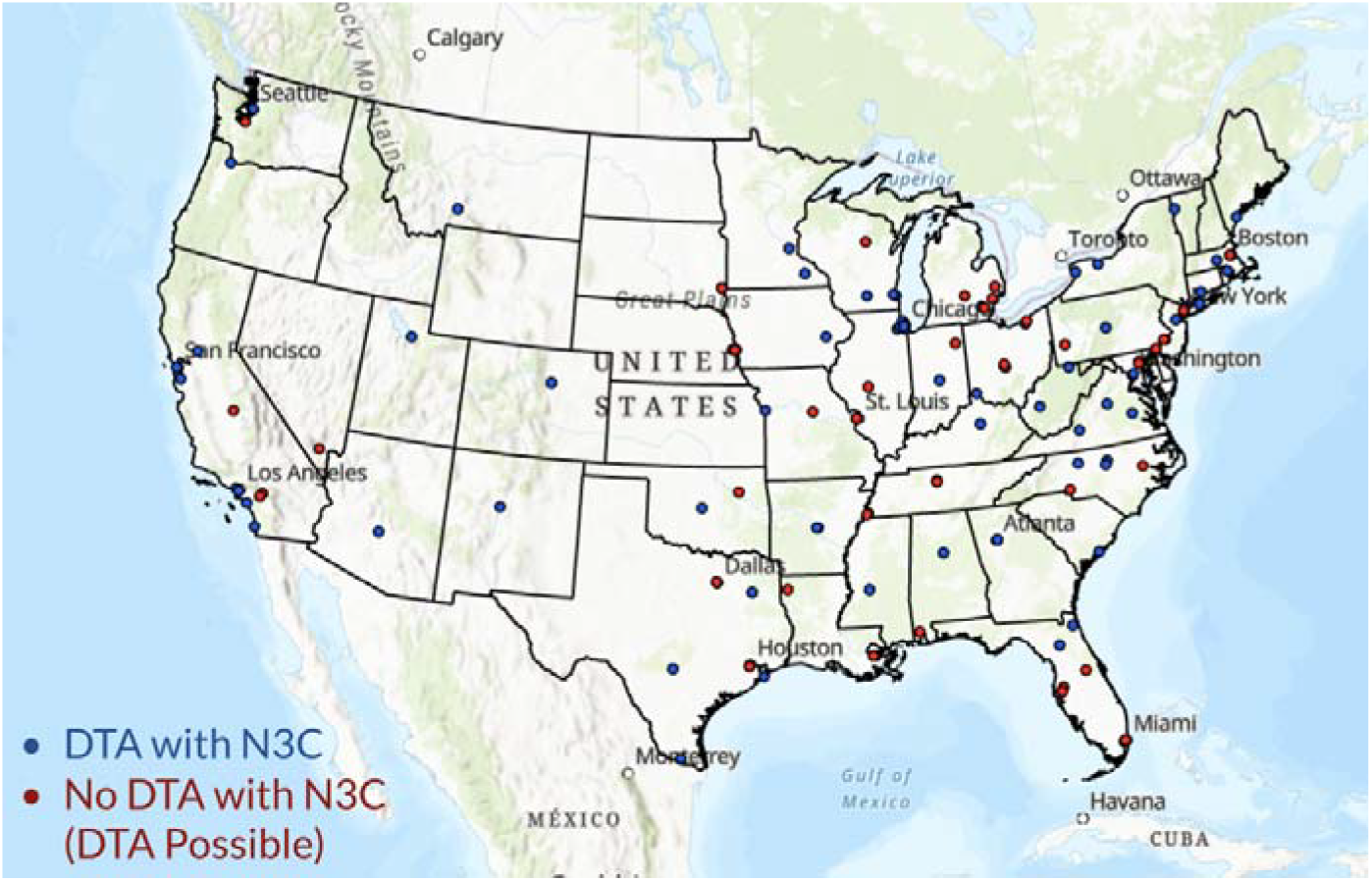
Distribution of N3C data contributors (blue) and institutions not contributing data but with data available to share (red). ArcGIS Pro 3.0.0 was used to generate this figure.

To test if institutional characteristics were associated with data sharing, we first grouped institutions as those that had data to share, defined as N3C participating institutions with hospital associations and the 11 large hospitals on Becker’s Hospital Review 2023 not participating in N3C. From the 137 institutions with clinical data to share, we defined the outcome of interest as whether the institution had a DTA (n=44 or 32% no). In univariate analyses, current NCATS funding (OR=4.09; 95% CI: 1.47-11.40; p=0.007) and at least one active FACMI (OR = 5.30; 95% CI: 1.51-7.87, p=0.009) were associated with data sharing among institutions with EHRs, whereas having a DBMI did not associate (Table 2). As a sensitivity analysis, institutions with a DBMI or a variant of a DBMI (centers or institutes) were associated with data sharing (OR = 4.22; 95% CI: 1.77-10.04; p=0.001), while the number of FACMI per institution did not associate (Table 2).

**Table 2.** Characteristics associated with N3C data sharing for institutions with data to share: Logistic regression univariate results for institutions that had data to share with N3C (n=137). The “Has DTA” column was calculated by filtering the 137 institutions to include just those with NCATS funding (n=37), an affiliated DBMI (n=17), or at least one FACMI (n=29) to determine the denominator for each of the independent variables. “Has DTA” column displays counts (percentages) or mean (standard deviation), where appropriate. Abbreviations: confidence interval (CI), department of biomedical informatics (DBMI), data transfer agreement (DTA), Fellows of the American College of Medical Informatics (FACMI), National Center for Advancing Translational Sciences (NCATS), odds ratio (OR).

|  | Has DTA | OR<br>(95% CI) | P-value |
| --- | --- | --- | --- |
| **Current NCATS Funding** | 32/37 (86.5%) | 4.09<br>(1.47-11.40) | 0.007 |
| **An Affiliated DBMI** | 14/17 (82.4%) | 2.42<br>(0.66-8.91) | 0.183 |
| **An Affiliated DBMI or Variant**<br>**(Centers and Institutes)** | 45/53 (84.9%) | 4.22<br>(1.77-10.04) | 0.001 |
| **At Least One FACMI** | 26/29 (89.7%) | 5.30<br>(1.51-18.63) | 0.009 |
| **Number of FACMI per**<br>**Institution** | 1.20 (3.12) | 1.34<br>(0.95-1.87) | 0.093 |

We also conducted a series of multivariate analyses. In the original model (Model 1), which included NCATS funding, an affiliated DBMI, and at least one active FACMI, no variable was significantly associated with data sharing while adjusting for the other two variables (Figure 3). However, when variants of DBMIs (centers and institutes) as opposed to departments only were included in the model (Model 2), this variable was associated with data sharing when adjusted for current NCATS funding and at least one active FACMI was associated with data sharing (OR = 3.03, 95% CI: 1.22-7.52; p=0.017). This association was slightly stronger when number of FACMI per institution were included as opposed to at least one FACMI (Model 3; OR = 3.28; 95% CI: 1.34-8.06; p=0.010). Lastly, current NCATS funding was significantly associated with data sharing when adjusted for an affiliated DBMI and the number of FACMI per institution (Model 4; OR = 3.19; 95% CI: 1.06-9.60; p=0.039).

**Figure 3:**
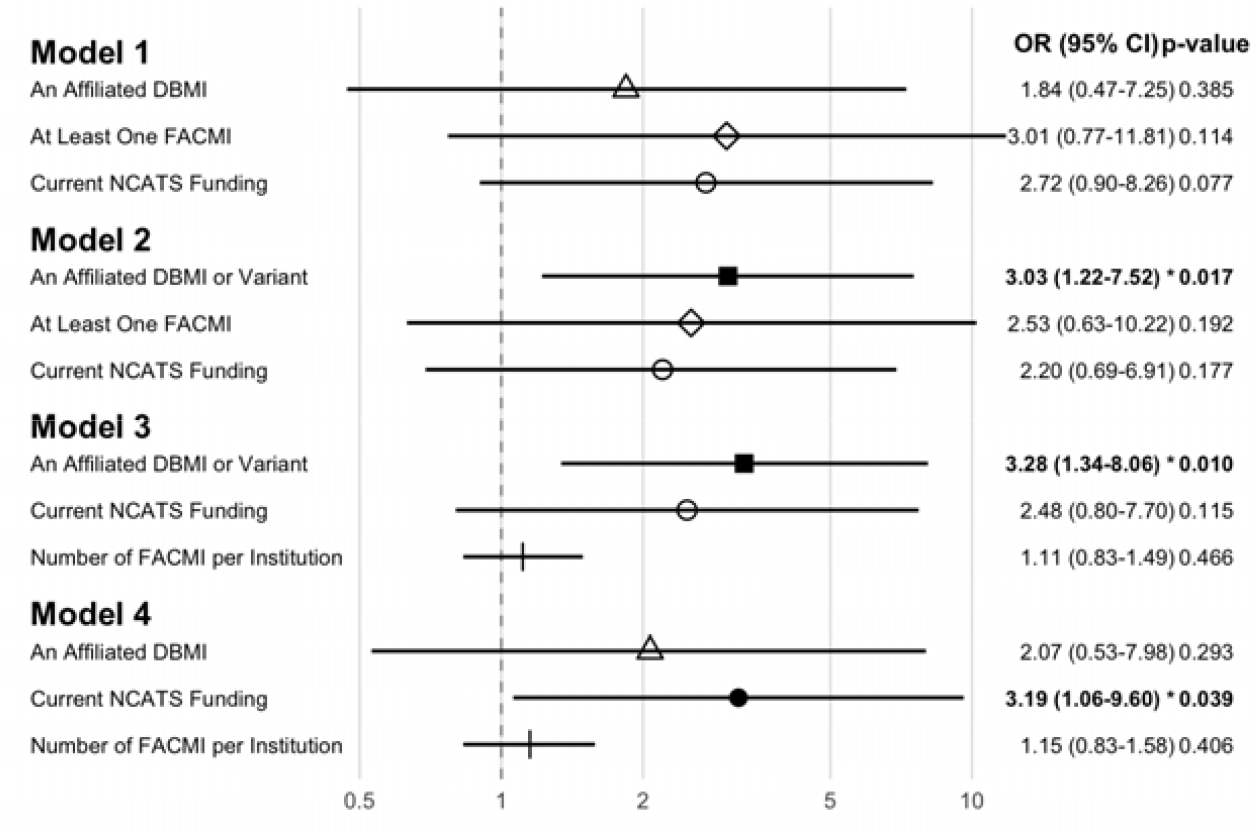
Characteristics associated with N3C data sharing for institutions with data to share: Logistic regression multivariate results for institutions that had data to share with N3C (n=137). Model 1 includes the Current NCATS Funding, Affiliated DBMI, and At Least One FACMI variables, Model 2 includes Current NCATS Funding, Affiliated DBMI or Variant, and At Least One FACMI variables, Model 3 includes Current NCATS funding, An Affiliated DBMI or Variant, and the number of FACMI per institution variables, and Model 4 includes Current NCATS funding, an Affiliated DBMI, and the number of FACMI per institution variables. Bolded values represent significance at the p<0.05 level. Abbreviations: confidence interval (CI), department of biomedical informatics (DBMI), Fellows of the American College of Medical Informatics (FACMI), National Center for Advancing Translational Sciences (NCATS), odds ratio (OR). R 4.4.3 was used to generate this figure.

Data sharing decisions at the institutional level are not well documented in the literature and consequently difficult to study or discuss based on empirical evidence. We hypothesize that some institutions might have been willing to share EHR data with N3C but instead made COVID-related research commitments with other groups. To determine if there were data to support this speculation, we gathered additional information about the institutions that had data available to share but did not have an executed DTA with N3C in place (n=44). We searched for evidence that these institutions were 1) contributing data to a COVID-19 registry outside of N3C, 2) enrolling participants in COVID-19 studies such as NIH’s Research COVID to Enhance Recovery (RECOVER^7,8^), or 3) engaged in local COVID-19 research, requiring access to institutional EHR data. We examined institutional webpages, searched PubMed by institution for published COVID-related research (excluding clinical trials and surveys among health professionals), and searched the internet for lists of COVID-19 research registry contributors. Contributing to a COVID-19 registry outside of the N3C included, but was not limited to, the American Heart Association (AHA) COVID-19 CVD registry^9^, the COVID-19 and Cancer Consortium (CCC19)^10^, and the Consortium for Clinical Characterization of COVID-19 by EHR (4CE)^11^. We found that 21 out of 44 (47.7%) of these institutions contributed to COVID-19-related registries or consortia but did not contribute to N3C. Additionally, 22 (50.0%) of these institutions were actively enrolling participants in COVID-19 related studies and 15 (34.1%) were actively engaged in COVID-related research requiring EHR access as of mid-2022. These data indicate that one-third to one-half of these institutions were not opposed to sharing or using EHR data for COVID-related research per se but that they simply chose not to share with N3C.

## Discussion

The emergence of SARS-CoV-2 and the resulting COVID-19 pandemic motivated one of the largest, rapid exchanges of US EHR data for research. While the fast-moving public health emergency was met with many willing partners, not all institutions embraced the accessible, yet secure data enclave developed for N3C equally. It is not clear from the literature why specific large and well-resourced institutions refrained and continue to refrain from contributing data to N3C. To better understand factors that impacted EHR data sharing during a public health emergency, we accessed public data to describe the institutional characteristics of institutions that shared data with N3C compared to those that did not but had the capacity to do so. Of the factors examined here, current NCATS funding, having affiliated departments, centers, or institutes of biomedical informatics, and active FACMI faculty were strongly associated with N3C data sharing in univariate tests of association. Having affiliated departments, centers, or institutes of biomedical informatics remained associated with data sharing in models that accounted for current NCATS funding and FACMI faculty.

While this study identified specific institutional characteristics associated with N3C data sharing, there are likely other characteristics not represented in this dataset associated with institutional data sharing decisions. Our observational study was limited to 137 institutions with EHR data to share with N3C as of mid-2022. The sample size is small and only powered to detect very large effect sizes in either unadjusted or adjusted analyses. The sample size has not appreciably increased since the mid-2022 data freeze, and this is not expected to change given the national COVID-19 public health emergency expired May 11, 2023. Based on these data, current NCATS funding, having affiliated departments, centers, or institutes of biomedical informatics, and active FACMI may represent an institution’s knowledge of and participation in initiatives that actively share data, likely predisposing that same institution to participate in other initiatives like N3C.

The variables identified as associated with N3C data sharing represent tangible targets for institutional initiatives aiming to increase clinical data sharing for research purposes. NCATS funds the Clinical and Translational Science Awards (CTSA) Program, and its current program goals include the creation of a national resource for rapid response to public health emergencies or needs (e.g., N3C). Per the latest Notice of Funding Opportunity PAR-24-272^12^, the NCATS CTSA awardees are expected to have “well-integrated and developed informatics and digital health capabilities.” NIH funding also comes with the expectation that data would be shared with the caveat that prescribed data sharing is limited to specific data types such as genomics^13^. We interpret current NCATS funding in this study as representative of the resources (funding, personnel, and infrastructure) that enable data sharing with N3C, indicating that increased NIH funding earmarked for informatics and data sharing would further increase data sharing. Increased NIH funding would, in turn, enable institutions to realize a partial return-of-investment for the substantial institutional investments made in infrastructure and expertise that are required as prerequisites to data sharing and research capacity^14^. Likewise, both NIH and individual institutions could target training programs and other points along the professional development pipeline to strengthen the biomedical and health informatics workforce and increase the number of capable faculty and staff.

Although our findings point to addressable variables that could increase data sharing with data enclaves such as N3C, we cannot fully explain why there are institutions with current NCATS funding, affiliated DBMIs and variants, and active FACMI that do not share data with N3C despite their knowledge of and participating in other COVID-19 data sharing activities. With the significant findings that DBMIs and variants were associated with data sharing with the N3C and the fact that the onboarding process for the N3C involves meetings with NIH officials, including data and hospital security personnel and IRB representatives, it is likely that these institutions who have been sharing data with the N3C have personnel who are well-versed in the protections of protected health information (PHI) for research applications and systems, as opposed to operational electronic health record systems (e.g., Epic EMR). Our limited findings based on public-facing institutional data described here indicate that decisions to exchange data with N3C are complex and may be institution specific. Collecting more nuanced data related to investments in infrastructure and data sharing decisions using qualitative approaches such as surveys and semi-structured interviews with institutional officials has the potential to identify additional, relevant variables. Qualitative approaches, however, rely on active participation. The use of public-facing institutional data used here is arguably free of the non-response bias likely introduced by the very institutions that do not contribute data to N3C despite their capacity to do so.

A feature of N3C that may have contributed to data sharing reservations is related to its choice of data science platforms. Specifically, the N3C Enclave includes Palantir Foundry^1^, a data science platform for data integration and data analysis developed by Palantir Technologies. Established in 2003, Palantir Technologies is a public company initially focused on the security market, with clients such as the US Army, Homeland Security, and Immigration and Customs Enforcement^15,16^. With the COVID-19 pandemic, Palantir Technologies expanded into the healthcare market with clients including the Centers for Disease Control and Prevention, the National Institutes of Health, and the National Health Service (NHS) England^15,16^. While no literature exists on reactions to NIH or CDC Palantir contracts, several reports, editorials, and letters voicing concerns ranging from data privacy to politics have been published in reaction to the pending NHS England contract with Palantir^16-21^. It may be that US institutions that did not contribute data to N3C have similar concerns but have not formally or publicly voiced them.

N3C, initially created specifically for COVID-19 research, is not the only research resource of aggregated EHRs (Table 3). Other general, aggregated EHR collections available for research include Epic Cosmos^22^ and TriNetX^23^. Cosmos was developed in collaboration with a community of health systems using Epic as their EHR vendor, with the aim of enhancing patient care^22^. Of the 44 institutions identified here that did not contribute data to N3C, 11 (27.5%) contributed data to Cosmos. Unlike N3C, the current limited dataset in the Cosmos Community does not include an analysis platform like Palantir Foundry, and while this may alleviate concerns related to third-party data misuse, the lack of a comparable analytics platform limits this version of Cosmos to basic count queries and visualizations^22^. The Cosmos Data Science Virtual Machine allows analysis of individual-level, de-identified data in an environment equipped with standard analysis tools (e.g., R, Python) but, unlike the NCATS-supported N3C, its use in sponsored research will incur as-of-yet unknown costs to be paid by the investigator. TriNetX, a commercial federated network^23^, was developed for patient cohort identification (primarily for clinical trials) and study feasibility counts, and it boasts an in-house analytics platform accessible via a web-portal. Unlike Cosmos, TriNetX is not limited to Epic EHR data and does not disclose the number of institutions or the specific institutions it includes. However, like Cosmos, TriNetX use may incur substantial fees depending on the research requests. While both Cosmos and TriNetX are larger than N3C, both are proprietary systems. With its mapping and harmonizing efforts along with data governance well documented in the public domain^24,25^, N3C distinguishes itself as an enclave developed to adhere to Findable, Accessible, Interoperable, and Reusable (FAIR) Principles^25,26^ and for reproducible research^1^.

**Table 3.** Characteristics of three large resources of aggregated clinical data for research. Abbreviations: electronic health record (EHR), graphical user interface (GUI), National Center for Advancing Translation Sciences (NCATS), National Clinical Cohort Collaborative (N3C), Observational Medical Outcomes Partnership (OMOP), structured query language (SQL).

|  | **NCATS N3C** | **Epic Cosmos** | **TriNetX** |
| --- | --- | --- | --- |
| **Type of sponsor** | Federal | Commercial | Commercial |
| **Data source** | Data source limited to US institutions regardless of EHR vendor entering into formal data transfer agreements | Data source limited to US institutions using Epic as the EHR vendor | Data source includes multiple US and non-US EHRs regardless of vendor and claims data |
| **User affiliation** | Member of institution with executed N3C data use agreement | Member of participating institution | Member of participating institution |
| **Data model** | OMOP | Epic (native) | OMOP |
| **Data de-duplication** | De-duplication based on privacy-preserving record linkage (PPRL), governed by a linkage honest broker agreement. | De-duplication based on Epic's CareEverywhere clinical health information exchange | No de-duplication |
| **Analysis platform** | Palantir Foundry | SlicerDicer (self-service cohort exploration tool); Data Science Virtual Machine (DSVM) | Web-based graphical analysis platform |
| **Access point** | Secure cloud enclave | Within Epic EHR | Web browser |
| **Interface type** | Code-based (Python/R) + visual tools | GUI + SQL | GUI + statistical tools |
| **Built-in statistics** | Custom and highly flexible | Limited | Propensity matching, Kaplan Meier |
| **Costs to user** | Free (federally funded) | SlicerDicer aggregate-level data access is generally free, but access flows through the user's institution's Epic relationship, and funded studies trigger a separate fee negotiation with Epic. DSVM is a paid access tier, requiring training certification (\$400–\$800) and additional Epic charge-backs for funded projects. Estimated project costs are unknown. | Self-service query and analytics platform are generally free for users at member institutions doing unfunded or internally funded work. Costs are negotiated and incurred when external grant funding is involved or when patient-level data extract is needed. Estimated project costs are unknown. |

The N3C and NCATS are piloting the use of the N3C infrastructure, initially developed for COVID-19 research, for other health conditions^27^. These pilot projects build on and extend N3C’s expertise and the public resources created in response to the pandemic. Each pilot project has its own data enclave, separate from the COVID Enclave, requiring distinct data transfer and use agreements^27^. Currently, there are two active pilot projects: the renal and cancer enclaves^27^. With the renewal of the COVID Enclave and expansion to pilot projects, understanding barriers that impact institutional sharing of clinical data for reproducible research will be key to optimizing this collaborative model and thereby enhancing research.

## Conclusion

Data access and data sharing, however complex and resource requiring, are essential for accurate and generalizable translational and policy-driving research. The COVID-19 public health emergency demonstrated the feasibility and power of real-world clinical data but also revealed a chronic reluctance of some US institutions to share data for the benefit of the larger general population. While it is not completely clear from public data alone why specific institutions did not share data during the pandemic, we found that institutions with departments, centers, or institutes of Biomedical Informatics were more likely to share data with the N3C, highlighting a potential pathway for improving data sharing initiatives in the future. However, it is clear that data or information not generally public may be key to further understanding the decisions to share for research purposes. US clinical data, already fragmented by its lack of a national healthcare system, will not reach its full research potential until barriers to data access and data sharing are identified, acknowledged, and completely dismantled.

## Data Availability

All data produced in the present study are available upon reasonable request to the authors

## Acknowledgements

This project was supported by the Clinical and Translational Science Collaborative of Northern Ohio, which is funded by the, UM1TR004528. The content is solely the responsibility of the authors and does not necessarily represent the official views of the NIH. CMR is supported by T32AG071474.

